# Immunohistochemical Detection of early and late Human Papillomaviruses (HPV) proteins in Retinoblastoma tumors

**DOI:** 10.1101/2020.07.16.20153882

**Authors:** Nara Diniz, Thatiana C Melo, Rodrigo P Araldi, Rodrigo F. Carvalho, Willy Beçak, Jonathan W de Medeiros, Giwellington S Albuquerque, Ana Pavla Gurgel, Rocio Hassan, Maria TC Muniz, Rita de Cassia Stocco

## Abstract

In this study, we evaluated the presence of early and late Human Papillomavirus (HPV) proteins in retinoblastoma Brazilian patients. For this, 8 formalin-fixed paraffin-embedded retinoblastoma tissue blocks were used. HPV DNA presence was determined by *in situ* hybridization (ISH). Immunohistochemistry were performed to verify HPV16/18 E6, E1^E4, and L1 proteins. HPV was detected in all retinoblastoma tumors and viral DNA was labeled in tumor cells, retinal layers and optical nerve structures. In addition, E1^E4, E6 and L1 proteins were detected in all samples in the same areas where HPV DNA was detected. Our data showed the presence and expression of early and late HPV proteins in retinoblastoma tumors from Brazilian children. However, further studies should be performed to clarify the role of HPV infection in retinoblastoma tumor.

## Introduction

Retinoblastoma is an intraocular tumor involving neuroectodermal cells of the retina, affecting 18,000-30,000 children per year (Antoneli et al., 2011; Chauhan et al., 2020; Orjuela et al., 2000; Palazzi et al., 2003; Ryoo et al., 2013; Sachdeva and O’Brien, 2012). Most retinoblastomas arise from inherited or acquired mutations in the *RB1* gene, leading to biallelic inactivation of this gene (Friend et al., 1986; Knudson, 1971). Tumors with intact *RB1* gene suggest other routes for their development. In this context, studies have demonstrated that HPV may play a role in the RB development since the E7 oncoprotein can inhibit action of pRb, destabilizing the cell cycle (Buitrago-Pérez et al., 2009; Dyson et al., 1989; Moody and Laimins, 2010; Orjuela et al., 2000; Whyte et al., 1988).

The prevalence of HPV in retinoblastoma has been shown with wide variations in different populations (4.6% - 69.7%) with the use of molecular methods for viruses detection (Antoneli et al., 2011; Bhuvaneswari et al., 2012; Espinoza et al., 2005; Gillison et al., 2007; Javanmard et al., 2019; Mohan et al., 2009; Montoya-Fuentes et al., 2003; Orjuela et al., 2000; Palazzi et al., 2003; Shetty et al., 2012; Shukla et al., 2009). Additionally, techniques such as *in situ* hybridization and immunohistochemistry have been performed and allowed possible causal relationship between HPV and RB development (Antoneli et al., 2011; Bhuvaneswari et al., 2012; Espinoza et al., 2005; Gillison et al., 2007; Palazzi et al., 2003; Shetty et al., 2012).

Although this relationship is not always suggested (Palazzi et al., 2003; Ryoo et al., 2013), child exposure to HPV during pregnancy and/or childbirth, indicates vertical viral transmission. In this scenario, it is suggested that HPV could be a factor in the development of oncogenic processes, supporting the idea that the virus is related, in fact, with the etiopathogenesis of RB (Bhuvaneswari et al., 2012; Lee et al., 2013; Skoczyński et al., 2014).

Evaluating the role of HPV in the development of retinoblastoma, we aimed to assess not only the viral presence in the tumor, but also possible viral activity, through detection of viral proteins in both retina and adjacent ocular tissues.

## Material and Methods

### Selected cases

The patients in this study were children diagnosed with retinoblastoma and treated at the “Center for Pediatric Oncohematology - University Hospital Oswaldo Cruz”. Eight formalin-fixed paraffin-embedded were obtained by spontaneous demand. Parents or guardians consented to the release of the material for research in retinoblastoma although data on eventual evaluations of the patient relatives were not included in this report. This study was performed with approval of the ethics committee of the Oswaldo Cruz University Hospital (CAEE0015.0.000-11).

### *In situ* hybridization

For the detection and typing of HPV in paraffin-embedded material, the *in situ* hybridization (ISH) technique was used. The detection was performed using the ZytoFast PLUS CISH Implementation Kit HRP-DAB (ZytoVision) with the ZytoFast HPV type 16/18/31/33/35 Probe (ZytoVision, T-1040-400), which identifies the 16, 18, 31, 33 and 35 types, according to the manufacturer’s instructions.

### Immunohistochemistry

The Immunohistochemistry technique was performed for the 8 paraffin-embedded cases, using EnVision ™ + system-HRP (AEC) (Dako) with antibodies (Abcam, AbcamInc, Cambridge, London) against HPV 16 E1 ^ E4 proteins (TV G402), HPV 1, 6, 11, 16, 18 and 31 L1 protein (ab2417) and HPV 16 and 18 E6 proteins (ab70).

## Results

RB tumor samples were analyzed for HPV presence by using *in situ* hybridization. HPV DNA was detected in all samples (n=8). The labeled areas included tumor cells, retinal layers and optical nerve structures (Figure 1 A, C and E).

**Figure 1.**
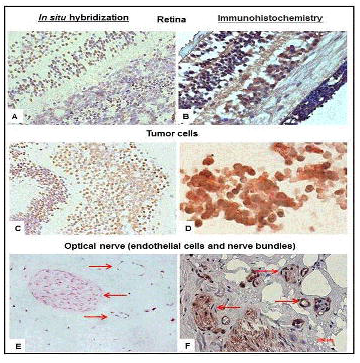
Comparative results of in hybridization and immunostaining for HPV in retinoblastoma. using the Green and DAB chromogens. A, C and E: Tumor cells hybridization in retinal areas, tumor and optic nerve, respectively. B, D and F: immunostaining in retinal areas (E6), tumer (E 1 *54) and optic Optical nerve (endothelial cells and nerve bundles) nerve (L1), respectively. Arrows in E and F.

To analyze the possible HPV activity, viral proteins were analyzed by immunohistochemistry. E1^E4, E6 and L1 proteins were visualized in retinoblastoma samples in the same areas where HPV DNA was detected (Figure 1B, D and F). These results showed early and late HPV genes expression.

## Discussion

The HPV relationship with the development of RB remains dubious, despite the detection of high and low risk HPV sequences have been detected in tumor samples (Antoneli et al., 2011; Shetty et al., 2012). In addition, HPV has been detected in tumor segments, ocular surface squamous indicating that the virus may be a risk factor for the development of other ocular tumors (Carreira et al., 2013).

Although Gillison et al. (2007) (Gillison et al., 2007) have not reported the presence of HPV in RB samples, suggesting that the virus has no role in the development of these tumors, we have shown the presence of high-risk HPV. High risk types have oncogenic potential by their oncoproteins ability to bind to the tumor suppressor proteins such as p53 and pRb, destabilizing the cell cycle and preventing apoptosis (McLaughlin-Drubin and Munger, 2009; Roman and Munger, 2013).

Unlike Ryoo et al. (2013) (Ryoo et al., 2013), which did not detected no viral DNA using *in situ* hybridization, the use of this technique allowed us to check the exact location of DNA in biological samples. The labeled areas included tumor cells, retinal layers and optical nerve structures. In addition, immunohistochemistry was performed to verify the presence of viral proteins in retinoblastoma samples. Thus, it was possible to verify the presence of DNA and proteins not only in tumor cells but also in adjacent tissues to the tumor. It was observed that the viral DNA is maintained in tumor tissues in advanced stages, reaching in some cases the optic nerve, with expression of important viral proteins, proteins that are related to the deregulation of the cell cycle (zur Hausen, 2002).

We have identified viral DNA and proteins in nerve bundles of optic nerve. Johnson et al. (2007) (Johnson et al., 2007) reported that retinoblastoma cells are subjected to an intensive neuronal differentiation in early stages. Therefore, according to these authors, the association between the presence of HPV and the morphological differentiation of this tumor suggests a possible role of HPV in the early stages of the RB development. Considering that the optic nerve arises from embryonic cells of the retina, the results are relevant due to the localization of virus and its proteins in verifying the viral location and activity in tumor development. In according, the identification of viral DNA was confirmed in fresh tumor.

In order to prove the presence and viral activity, we analyzed three different HPV proteins, that demonstrated that HPV is not only present in the samples analyzed as well as expressing its protein. The analysis of viral activity, targeting E1^E4, E6 and L1 proteins in retinoblastoma samples, showed that these proteins could be detected in the same areas where HPV DNA was detected. The identification of HPV DNA and L1 protein in the same regions of the eye tissue suggests this tissue as an organization site for productive infection. In addition, proteins required for viral replication (E1 ^ E4) were detected in the other regions, suggesting viral DNA replication. The presence of the immunolabeled E6 protein also indicates the transforming action of HPV in cells, since this oncoprotein can bind and degrade the p53 protein (McLaughlin-Drubin and Munger, 2009). More studies are needed to identify other proteins such as L2, E2 and E7, to verify the possible complete formation of the viral particle, its integralization to the host genome and its transforming action on this tumor stage and in these locations.

It is well described in the literature that the HPV complete its cycle in differentiated, epithelial, cells (McLaughlin-Drubin and Munger, 2009) and expression of structural genesis is restricted to keratinocytes. Our results suggest that the expression of viral proteins cannot depend solely on differentiation of keratinocytes. We have identified the HPV proteins in tumor cells, different layers of the retina, cornea epithelial tissue and nerve cells constituting the optic nerve, as well as in inflammatory infiltrates.

Viral sources in these young individuals should be discussed. It is suggested that HPV infects the child during childbirth (Bhuvaneswari et al., 2012), or through vertical transmission, since HPV has been detected in the peripheral blood of pregnant women, umbilical cord of newborns (Tseng et al., 1992) and amniotic fluid (Armbruster-Moraes et al., 1994; Xu et al., 1998). Therefore, the virus can induce retinoblastoma and other oncogenic processes (Tseng et al., 1992). In addition, HPV has been detected in trophoblasts, through which nutrient exchange occurs between the mother and the fetus. This infection has been reported as the cause of some miscarriages (Hermonat et al., 1998).

Since the origin of HPV infection in children with retinoblastoma may be through vertical transmission (Bhuvaneswari et al., 2012), reducing the incidence of this virus in women may lead to a decrease in cancers linked to viral infection. Efforts to reduce cases of HPV infection, lesions and/or cervical cancer are extremely important for the reduction of cases of sporadic retinoblastomas involving HPV. Like cervical cancer, the reduction of RB cases indirectly by the monitoring of women, causes a decrease in the traumatic processes of the children in their treatment, which often includes the surgical process of enucleation, as well as chemotherapy and radiotherapy.

## Conclusions

The results of this study showed the presence of HPV DNA in retinoblastoma tumors. In addition, early and late HPV proteins were detected in all samples, suggesting a viral activity in retinoblastoma tumors.

## Data Availability

The authors confirm that the data supporting the findings of this study are available within the article.

## Competing Interests

The authors declare that they do not have anything to disclose regarding funding from industries or conflict of interest with respect to this manuscript.

## Authors’ Contributions

ND and JWM carried out the molecular genetic studies, hybridization assays, and drafted the manuscript. TCM and GSA participed in immunoassays. RPA participated in hybridization assays. WB participated in the design of the study. MTCM, APG, RS and participated in the design of the study, and helped to draft the manuscript. RCS and conceived of the study, and participated in its design and coordination, RFC join RCS in revision. All authors read and approved the final manuscript.

## Acknowledgments

The authors acknowledge the CNPq (Conselho Nacional de DesenvolvimentoCientífico e Tecnológico), and CAPES (Coordenação de Aperfeiçoamento de Pessoal de Nível Superior) that supported this study.

## Notes

### Competing Interest Statement

The authors have declared no competing interest.

### Funding Statement

The authors acknowledge the CNPq (Conselho Nacional de Desenvolvimento Cientifico e Tecnologico), and CAPES (Coordenacao de Aperfeicoamento de Pessoal de Nivel Superior) that supported this study.

### Author Declarations

This study was performed with approval of the ethics committee of the Oswaldo Cruz University Hospital (CAEE0015.0.000-11).

## References

Antoneli, C.B.G., Ribeiro, K.B., Sredni, S.T., Arias, V.E.A., Andreoli, M.A., de Camargo, B., Sobrinho, J.S., Prado, J.C.M., Soares, F.A., Villa, L.L., 2011. Low prevalence of HPV in Brazilian children with retinoblastoma. J. Med. Virol. 83, 115–118. https://doi.org/10.1002/jmv.21925

Armbruster-Moraes, E., Ioshimoto, L.M., Leão, E., Zugaib, M., 1994. Presence of Human Papillomavirus DNA in Amniotic Fluids of Pregnant Women with Cervical Lesions. Gynecol. Oncol. 54, 152–158. https://doi.org/10.1006/gyno.1994.1185

Bhuvaneswari, A., Pallavi, V.R., Jayshree, R.S., Kumar, R.V., 2012. Maternal transmission of human papillomavirus in retinoblastoma: A possible route of transfer. Indian J. Med. Paediatr. Oncol. Off. J. Indian Soc. Med. Paediatr. Oncol. 33, 210–215. https://doi.org/10.4103/0971-5851.107080

Buitrago-Pérez, Á., Garaulet, G., Vázquez-Carballo, A., Paramio, J.M., García-Escudero, R., 2009. Molecular Signature of HPV-Induced Carcinogenesis: pRb, p53 and Gene Expression Profiling. Curr. Genomics 10, 26–34. https://doi.org/10.2174/138920209787581235

Carreira, H., Coutinho, F., Carrilho, C., Lunet, N., 2013. HIV and HPV infections and ocular surface squamous neoplasia: systematic review and meta-analysis. Br. J. Cancer 109, 1981–1988. https://doi.org/10.1038/bjc.2013.539

Chauhan, S., Sen, S., Singh, N., Sharma, A., Chawla, B., Kashyap, S., 2020. Human Papillomavirus Detection Strategies in Retinoblastoma. Pathol. Oncol. Res. 26, 1341–1344. https://doi.org/10.1007/s12253-018-00577-x

Dyson, N., Howley, P.M., Münger, K., Harlow, E., 1989. The human papilloma virus-16 E7 oncoprotein is able to bind to the retinoblastoma gene product. Science 243, 934–937. https://doi.org/10.1126/science.2537532

Espinoza, J.P.M., Cardenas, V.J.P., Luna, C.A., Fuentes, H.M., Camacho, G.V., Carrera, F.M., Garcia, J.R.G., 2005. Loss of 10p material in a child with human papillomavirus–positive disseminated bilateral retinoblastoma. Cancer Genet. Cytogenet. 161, 146–150. https://doi.org/10.1016/j.cancergencyto.2005.01.012

Friend, S.H., Bernards, R., Rogelj, S., Weinberg, R.A., Rapaport, J.M., Albert, D.M., Dryja, T.P., 1986. A human DNA segment with properties of the gene that predisposes to retinoblastoma and osteosarcoma. Nature 323, 643–646. https://doi.org/10.1038/323643a0

Gillison, M.L., Chen, R., Goshu, E., Rushlow, D., Chen, N., Banister, C., Creek, K.E., Gallie, B.L., 2007. Human retinoblastoma is not caused by known pRb-inactivating human DNA tumor viruses. Int. J. Cancer 120, 1482–1490. https://doi.org/10.1002/ijc.22516

Hermonat, P.L., Kechelava, S., Lowery, C.L., Korourian, S., 1998. Trophoblasts are the preferential target for human papilloma virus infection in spontaneously aborted products of conception. Hum. Pathol. 29, 170–174. https://doi.org/10.1016/s0046-8177(98)90228-3

Javanmard, D., Moein, M., Esghaei, M., Naseripour, M., Monavari, S.H., Bokharaei-Salim, F., Sadeghipour, A., 2019. Molecular evidence of human papillomaviruses in the retinoblastoma tumor. VirusDisease 30, 360–366. https://doi.org/10.1007/s13337-019-00540-7

Johnson, D.A., Zhang, J., Frase, S., Wilson, M., Rodriguez-Galindo, C., Dyer, M.A., 2007. Neuronal Differentiation and Synaptogenesis in Retinoblastoma. Cancer Res. 67, 2701–2711. https://doi.org/10.1158/0008-5472.CAN-06-3754

Knudson, A.G., 1971. Mutation and Cancer: Statistical Study of Retinoblastoma. Proc. Natl. Acad. Sci. U. S. A. 68, 820–823.

Lee, S.M., Park, J.S., Norwitz, E.R., Koo, J.N., Oh, I.H., Park, J.W., Kim, S.M., Kim, Y.H., Park, C.-W., Song, Y.S., 2013. Risk of vertical transmission of human papillomavirus throughout pregnancy: a prospective study. PloS One 8, e66368. https://doi.org/10.1371/journal.pone.0066368

McLaughlin-Drubin, M.E., Munger, K., 2009. Oncogenic Activities of Human Papillomaviruses. Virus Res. 143, 195–208. https://doi.org/10.1016/j.virusres.2009.06.008

Mohan, A., Venkatesan, N., Kandalam, M., Pasricha, G., Acharya, P., Khetan, V., Gopal, L., Sharma, T., Biswas, J., Krishnakumar, S., 2009. Detection of human papillomavirus DNA in retinoblastoma samples: a preliminary study. J. Pediatr. Hematol. Oncol. 31, 8–13. https://doi.org/10.1097/MPH.0b013e31818b373b

Montoya-Fuentes, H., de la Paz Ramirez-Munoz, M., Villar-Calvo, V., Suarez-Rincon, A.E., Ornelas-Aguirre, J.M., Vazquez-Camacho, G., Orbach-Arbouys, S., Bravo-Cuellar, A., Sanchez-Corona, J., 2003. Identification of DNA sequences and viral proteins of 6 human papillomavirus types in retinoblastoma tissue. Anticancer Res. 23, 2853–2862.

Moody, C.A., Laimins, L.A., 2010. Human papillomavirus oncoproteins: pathways to transformation. Nat. Rev. Cancer 10, 550–560. https://doi.org/10.1038/nrc2886

Orjuela, M., Castaneda, V.P., Ridaura, C., Lecona, E., Leal, C., Abramson, D.H., Orlow, I., Gerald, W., Cordon-Cardo, C., 2000. Presence of human papilloma virus in tumor tissue from children with retinoblastoma: an alternative mechanism for tumor development. Clin. Cancer Res. Off. J. Am. Assoc. Cancer Res. 6, 4010–4016.

Palazzi, M.A., Yunes, J.A., Cardinalli, I.A., Stangenhaus, G.P., Brandalise, S.R., Ferreira, S.A., Sobrinho, J.S.P., Villa, L.L., 2003. Detection of oncogenic human papillomavirus in sporadic retinoblastoma. Acta Ophthalmol. Scand. 81, 396–398. https://doi.org/10.1034/j.1600-0420.2003.00112.x

Roman, A., Munger, K., 2013. The papillomavirus E7 proteins. Virology, Special Issue: The Papillomavirus Episteme 445, 138–168. https://doi.org/10.1016/j.virol.2013.04.013

Ryoo, N.-K., Kim, J.-E., Choung, H.-K., Kim, N., Lee, M.-J., Khwarg, S.-I., 2013. Human Papilloma Virus in Retinoblastoma Tissues from Korean Patients. Korean J. Ophthalmol. KJO 27, 368–371. https://doi.org/10.3341/kjo.2013.27.5.368

Sachdeva, U.M., O’Brien, J.M., 2012. Understanding pRb: toward the necessary development of targeted treatments for retinoblastoma. J. Clin. Invest. 122, 425–434. https://doi.org/10.1172/JCI57114

Shetty, O.A., Naresh, K.N., Banavali, S.D., Shet, T., Joshi, R., Qureshi, S., Mulherkar, R., Borges, A., Desai, S.B., 2012. Evidence for the presence of high risk human papillomavirus in retinoblastoma tissue from nonfamilial retinoblastoma in developing countries. Pediatr. Blood Cancer 58, 185–190. https://doi.org/10.1002/pbc.23346

Shukla, S., Bharti, A.C., Mahata, S., Hussain, S., Kumar, R., Hedau, S., Das, B.C., 2009. Infection of human papillomaviruses in cancers of different human organ sites. Indian J. Med. Res. 130, 222–233.

Skoczyński, M., Goździcka-Józefiak, A., Kwaśniewska, A., 2014. Risk factors of the vertical transmission of human papilloma virus in newborns from singleton pregnancy - preliminary report. J. Matern.-Fetal Neonatal Med. Off. J. Eur. Assoc. Perinat. Med. Fed. Asia Ocean. Perinat. Soc. Int. Soc. Perinat. Obstet. 27, 239–242. https://doi.org/10.3109/14767058.2013.807238

Tseng, C.J., Lin, C.Y., Wang, R.L., Chen, L.J., Chang, Y.L., Hsieh, T.T., Pao, C.C., 1992. Possible transplacental transmission of human papillomaviruses. Am. J. Obstet. Gynecol. 166, 35–40. https://doi.org/10.1016/0002-9378(92)91825-u

Whyte, P., Buchkovich, K.J., Horowitz, J.M., Friend, S.H., Raybuck, M., Weinberg, R.A., Harlow, E., 1988. Association between an oncogene and an anti-oncogene: the adenovirus E1A proteins bind to the retinoblastoma gene product. Nature 334, 124–129. https://doi.org/10.1038/334124a0

Xu, S., Liu, L., Lu, S., Ren, S., 1998. Clinical observation on vertical transmission of human papillomavirus. Chin. Med. Sci. J. Chung-Kuo Hsueh Ko Hsueh Tsa Chih 13, 29–31.

zur Hausen, H., 2002. Papillomaviruses and cancer: from basic studies to clinical application. Nat. Rev. Cancer 2, 342–350. https://doi.org/10.1038/nrc798

